# The gender gap in the relationship between Metabolic Syndrome and Restrictive Ventilatory Defect

**DOI:** 10.1101/2023.12.20.23300321

**Authors:** Ya-Chun Chu, Chi-Chiang Yang, Shaw-Ji Chen, Pei-Ling Cheng, Mei-Chuan Wu, Hsin-Hung Wu, Jerry Cheng-Yen Lai

**Author notes:** Corresponding Authors: **Hsin-Hung Wu, MD,** (HH Wu) **Jerry Cheng-Yen Lai, PhD,** (JC Lai).

## Abstract

**Background:** Given the fundamental physiological differences between the sexes, this study aimed to investigate the effect of metabolic syndrome on ventilatory defects stratified by sex.

**Methods:** We conducted a nationwide, pooled, cross-sectional study. Data of 45,788 participants (men, n=15,859; women, n=29,929) aged 30 years or more were obtained from the Taiwan Biobank. Age-sex adjusted and multivariate logistic regression models were used to estimate the risk of developing impaired pulmonary function (restrictive or obstructive ventilatory defect) in individuals with or without metabolic syndromes. Separate models were also used to estimate for the risk of metabolic syndrome scores and the risk of individual metabolic abnormalities on the risk of restrictive ventilatory defect.

**Results:** The overall prevalence of metabolic syndrome was estimated to be 15.9% in Taiwan, much higher in men than in women (18.6% versus 14.4%). Significant association was observed on the effect of metabolic syndromes on the risk of restrictive ventilatory defect. The risk of developing restrictive ventilator defect was 35% higher in participants with metabolic syndromes (odds ratio, 1.35; 95% confidence interval, 1.26-1.45) than those without metabolic syndromes. Elevated blood pressure and triglycerides abnormality were important predictors of restrictive ventilator defect.

Sex-stratified subgroup analyses of the individual metabolic abnormalities indicated that men with abdominal obesity and women with dysglycemia were more likely to develop restrictive ventilatory defect.

**Conclusion:** Our study evidences suggested that metabolic syndromes were important predictors of impaired pulmonary function and increased risk of developing restrictive ventilatory defects, and its risk increased with increasing numbers of metabolic abnormalities.

## Introduction

Metabolic syndrome (MetS) is a condition that includes a cluster of risk metabolic abnormalities for a variety of diseases, including abdominal obesity (increasing waist circumference [WC]), high blood pressure (BP), high level of fasting plasma glucose (FPG), high level of triglyceride (TG), and low level of high-density lipoprotein cholesterol (HDL-C) levels. The MetS prevalence rates differed considerably from as high as 39.4%-57.1% in several Western countries (Finland, Italy, Mexico) to as low as 3.2%-15.5% (Poland, China, Turkey) [1]. In contrast, MetS is one of the most rapidly increasing chronic conditions in the Taiwanese population. According to the Nutrition and Health Survey in Taiwan, the prevalence rate of MetS has nearly tripled from 11.9% in 1993 to 34.6% in 2020 in the adult population [2]. An important sequel is the increased risk of cardiovascular disease, diabetes, hypertension, and hyperlipidemia in people with MetS, which is 2 to 6 times higher than in normal people [3]. Inadequate pulmonary ventilation would irritate the medulla oblongata to compensate by rising respiratory rates. This situation may lower exercise tolerance or cause shortness of breath and orthopnea. Compensation failure would contribute to chronic hypoxia or muscle atrophy, and long-term respiratory failure could lead to changes in lung structure, increasing the risk of pulmonary hypertension and Cor Pulmonale [4]. Given the fundamental physiological differences between the sexes, women used more movement of the chest and ribs than men during deep breathing, while men mainly applied the diaphragm expansion and abdominal muscles to breathe [5]. The Korean National Health and Nutrition Examination Survey reported that WC, BP, FPG, and HDL-C independently related to forced vital capacity of predicted % (FVC % of predicted) in men while waist circumference significantly correlated with FVC predicted (%) in women [6]. In contrast, in a study of 6,945 Taiwanese participants, a negative correlation was reported between WC and FVC % predicted in men younger than 45 years, while no correlation between metabolic abnormality and FVC % predicted was observed in women [7].

Despite many published studies on the association between MetS and impaired pulmonary function, no consistent evidences are available in the literature by sex. We aimed to compare the risk of developing impaired pulmonary function between individuals with or without MetS for each gender using a nationwide, pooled, cross-sectional study design in Taiwan. We hypothesized that sex was an independent predictor of impaired pulmonary functions, and the effects of MetS on impaired pulmonary functions differed by sex among community-dwelling adult population in Taiwan.

## Methods

### 1. Study design and data source

We conducted a nationwide, pooled, cross-sectional study using the data obtained from the Taiwan Biobank. Taiwan Biobank is a largescale biomedical research database that intends to recruit 200,000 community-dwelling healthy volunteers aged 30-79 years residing in Taiwan that consists of genetic and medical information as on predominantly Han Chinese-ancestry individuals [8]. All participants in the Taiwan Biobank provided written informed consent before data collection. The vertical data integration of the collected samples provide data accessible to the community dwelling population in Taiwan. As stipulated by the Human Biobank Management Act, the data of all participants were de-identified with uniquely encrypted identification numbers in the Taiwan Biobank. As a reliable data source, studies using this database have been published in top-tier journals [9, 10]. This study has been reviewed and approved by the Human Research Ethics Review Committee of the MacKay Memorial Hospital (21MMHIS351e).

### 2. Study population

Using data released in 2021, a total of 132,720 adult participants recruited between 2008 and 2020 were available for analyses. The exclusion criteria were: (1) participants with missing information on pulmonary function (n=50,310) and metabolic abnormalities (n=47); (2) to ensure the precision of the blood glucose and lipids test results, blood sugar test results with less than 8 hours of fasting were excluded (n=21,854); (3) to maintain homogeneity in prognosis, we excluded all participants with cancers, respiratory diseases, cardiovascular diseases, and metabolic-related diseases (n=13,011), and covariates due to incomplete data (n=1,710). A total of 45,788 participants aged 30 years or more were included in the final analysis.

### 3. Analysis of covariates

Participants’ residential urbanicity was defined according to Liu et al [11]. Education levels were divided into college and above (university and graduate school or above), middle school education (junior high and senior high school) and elementary school (elementary school or illiterate). Participants were classified into never smoking, former smoking (did not smoke in recent six months), or current smoking (for more than six months). The habit of drinking was defined as 150cc per week for 6 months, and former drinking was defined as not drinking for more than 6 months. Monthly exercise habits classified the participants using the metabolic equivalent for task (MET) as never or seldom physical activity, doing light physical activity (monthly physical exercise <1800 MET), moderate physical activity (monthly physical exercise=1800-3600 MET), or vigorous physical activity (monthly physical exercise > 3600 MET) [12]. MET is a unit that estimates the amount of energy (kcal/kg/hour) used by an individual during physical activity, and the values were multiplied by 150 minutes as suggested by the World Health Organization [13].

### 4. Definition of metabolic abnormalities

According to the modified National Cholesterol Education Program Adult Treatment Panel III (NCEP ATP III), individuals having at least three of the five metabolic abnormalities were diagnosed as having MetS [14]: 1) abdominal obesity: WC ≥90 cm in men and ≥80 cm in women; 2) hypertriglyceridemia: TGs ≥ 150 mg/dL or pharmacotherapy for hyperlipidemia; 3) reduced HDL-C level: HDL-C < 40 mg/dL in men and < 50 mg/dL in women; 4) elevated BP: systolic blood pressure ≥130mmHg or diastolic blood pressure ≥ 85 mmHg or pharmacotherapy for hypertension; 5) elevated FPG ≥ 100 mg/dL or pharmacotherapy for glucose-lowering medication.

### 5. Pulmonary function testing

The pulmonary function was performed by Lilly-type pneumotach sensor spirometer (HI-801, CHEST M.I., INC, Tokyo, Japan) with specially trained technicians according to the American Thoracic Society recommendations. All spirometric parameters were repeated for three or more times with similar results to meet the criteria for acceptability and reproducibility. The spirometric parameters were compared with the estimated values of the prediction lung function formula adjusting for race, sex, age, and height, which were used to calculate FEV1 (forced expiratory volume in one second) predicted %, FVC (forced vital capacity) predicted % and FEV1 to FVC ratio. Respiratory dysfunction can be classified into obstructive ventilatory defect (OVD) and restrictive ventilatory defect (RVD). The pulmonary function test results were classified as normal ventilatory function (FEV1/FVC≧70%), RVD (FEV1/FVC≧70% & FVC<80% of predicted), and OVD (FEV1/FVC<70%) [15–18].

### 6. Statistical analysis

Chi-squared test for categorial variables and Student’s t-test for continuous variables were used to compare group differences. Age-sex adjusted and multivariate logistic regression models were used to estimate the risk of developing impaired pulmonary function (RVD or OVD) between individuals with or without MetS stratified by sex and age. Separate models were also used to estimate for the risks of metabolic syndrome scores (MSS) and individual metabolic abnormalities on the risk of RVD. The reported P values were based on a two-tailed test, and P <0.05 indicated a statistically significant. All data conversions and analyses were performed using SAS for Windows release 9.4 (SAS Institute Inc, Cary, NC).

## Results

### 1. Baseline characteristics

Table 1 compares basic demographic characteristic (age, obese status, residential urbanicity, and education level), healthy lifestyle behaviors, family medical history, metabolic abnormalities, and information related to pulmonary impairment between sexes. Most participants were women (65.4%) aged 30-59 years, resided in urbanized community, and attended college or graduate school. In general, men had a significantly higher incidences of metabolic abnormalities than women, including elevated FPG (29.3% vs 17.2%), TGs (28.8% vs 15.2%), and BP (36.8% vs 20.7%). On the contrary, women had higher incidences of abdominal obesity (28.3% vs 19.3%) and high-density cholesterol (26.8% vs 21.6%).

**Table 1.**
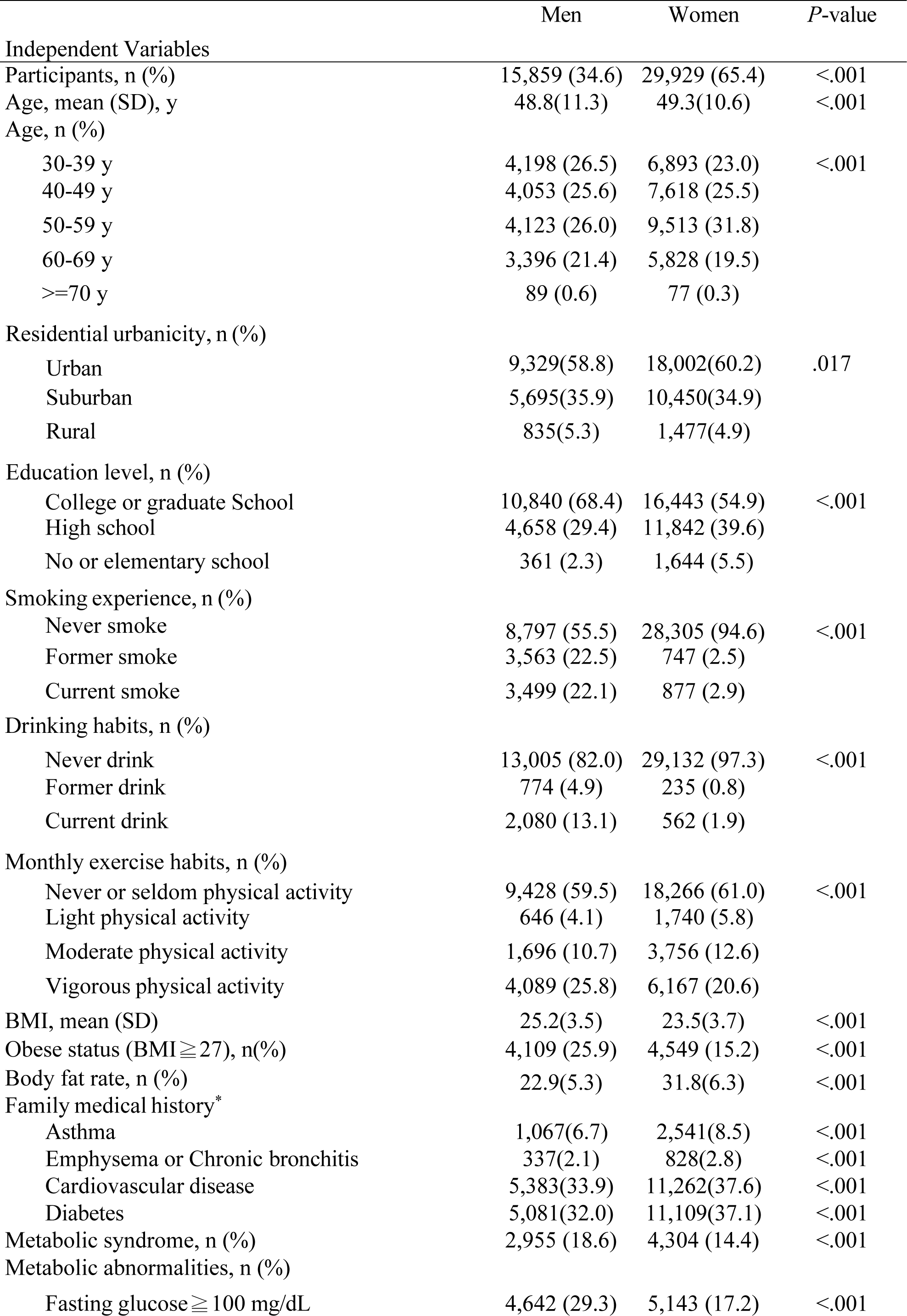
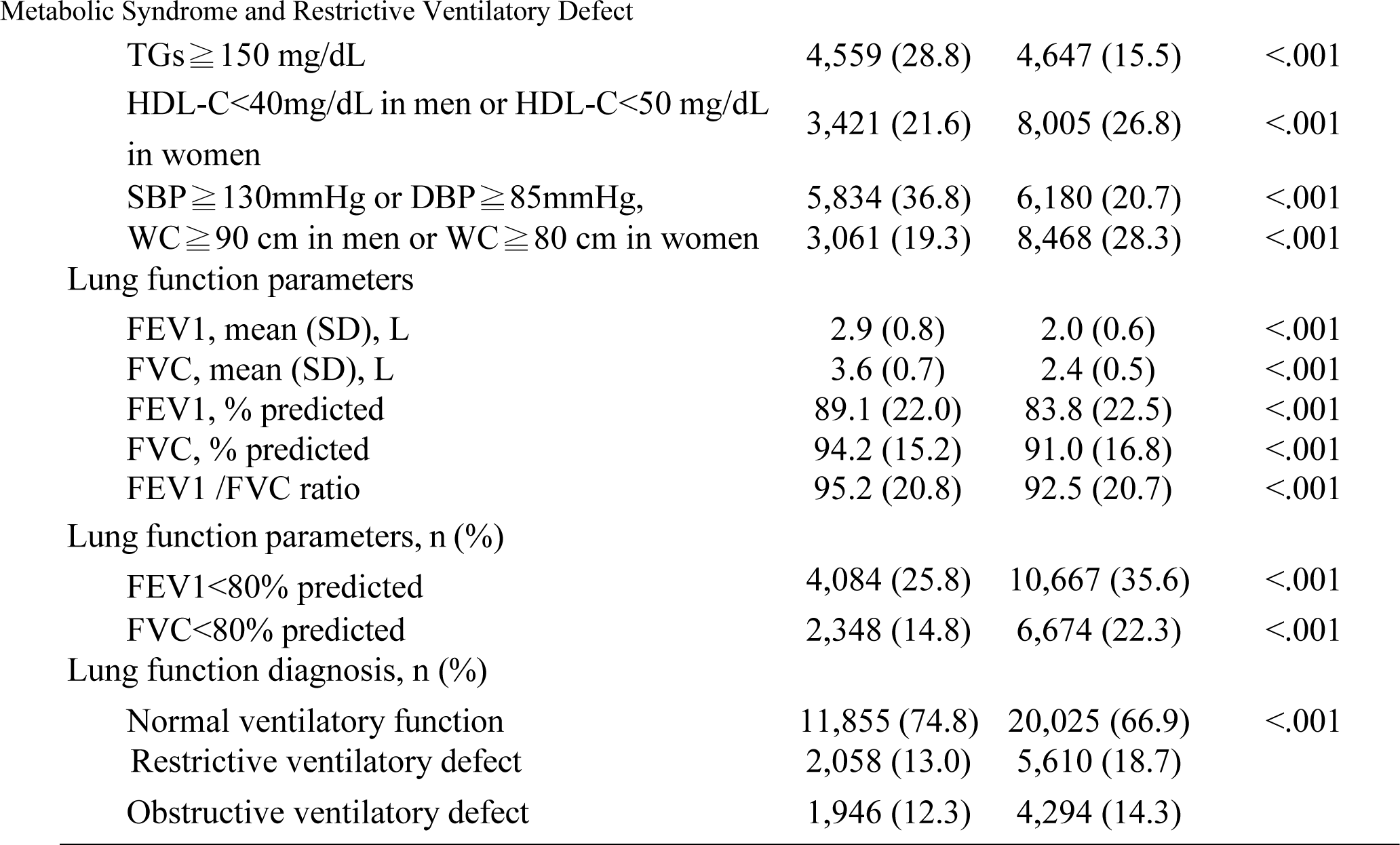
Baseline characteristics of total participants stratified by sex (N=45,788). SD, standard deviation; Mets, metabolic syndrome; WC, waist circumference; BMI, body mass index; SBP, systolic blood pressure; DBP, diastolic blood pressure; TGs, triglycerides; HDL-C, High-density lipoprotein Cholesterol; FVC, Forced volume capacity; FEV1,Force expiratory volume in one second; *P*-value, χ^2^ test or Student *t*-test. * Family medical history of the participant’s second degree of blood relatives were included in the family medical history. Cardiovascular disease consisted of vascular heart disease, coronary heart disease, arrhythmia, cardiomyopathy, congenital heart disease, hyperlipidemia, hypertension, and stroke.

|  | Men | Women | *P*-value |
| --- | --- | --- | --- |
| **Independent Variables** |  |  |  |
| Participants, n (%) | 15,859 (34.6) | 29,929 (65.4) | <.001 |
| Age, mean (SD), y | 48.8(11.3) | 49.3(10.6) | <.001 |
| Age, n (%) |  |  |  |
| 30-39 y | 4,198 (26.5) | 6,893 (23.0) | <.001 |
| 40-49 y | 4,053 (25.6) | 7,618 (25.5) |  |
| 50-59 y | 4,123 (26.0) | 9,513 (31.8) |  |
| 60-69 y | 3,396 (21.4) | 5,828 (19.5) |  |
| ≥70 y | 89 (0.6) | 77 (0.3) |  |
| Residential urbanicity, n (%) |  |  |  |
| Urban | 9,329(58.8) | 18,002(60.2) | .017 |
| Suburban | 5,695(35.9) | 10,450(34.9) |  |
| Rural | 835(5.3) | 1,477(4.9) |  |
| Education level, n (%) |  |  |  |
| College or graduate School | 10,840 (68.4) | 16,443 (54.9) | <.001 |
| High school | 4,658 (29.4) | 11,842 (39.6) |  |
| No or elementary school | 361 (2.3) | 1,644 (5.5) |  |
| Smoking experience, n (%) |  |  |  |
| Never smoke | 8,797 (55.5) | 28,305 (94.6) | <.001 |
| Former smoke | 3,563 (22.5) | 747 (2.5) |  |
| Current smoke | 3,499 (22.1) | 877 (2.9) |  |
| Drinking habits, n (%) |  |  |  |
| Never drink | 13,005 (82.0) | 29,132 (97.3) | <.001 |
| Former drink | 774 (4.9) | 235 (0.8) |  |
| Current drink | 2,080 (13.1) | 562 (1.9) |  |
| Monthly exercise habits, n (%) |  |  |  |
| Never or seldom physical activity | 9,428 (59.5) | 18,266 (61.0) | <.001 |
| Light physical activity | 646 (4.1) | 1,740 (5.8) |  |
| Moderate physical activity | 1,696 (10.7) | 3,756 (12.6) |  |
| Vigorous physical activity | 4,089 (25.8) | 6,167 (20.6) |  |
| BMI, mean (SD) | 25.2(3.5) | 23.5(3.7) | <.001 |
| Obese status (BMI ≥ 27), n(%) | 4,109 (25.9) | 4,549 (15.2) | <.001 |
| Body fat rate, n (%) | 22.9(5.3) | 31.8(6.3) | <.001 |
| Family medical history* |  |  |  |
| Asthma | 1,067(6.7) | 2,541(8.5) | <.001 |
| Emphysema or Chronic bronchitis | 337(2.1) | 828(2.8) | <.001 |
| Cardiovascular disease | 5,383(33.9) | 11,262(37.6) | <.001 |
| Diabetes | 5,081(32.0) | 11,109(37.1) | <.001 |
| Metabolic syndrome, n (%) | 2,955 (18.6) | 4,304 (14.4) | <.001 |
| Metabolic abnormalities, n (%) |  |  |  |
| Fasting glucose ≥ 100 mg/dL | 4,642 (29.3) | 5,143 (17.2) | <.001 |
| TGs ≥ 150 mg/dL | 4,559 (28.8) | 4,647 (15.5) | <.001 |
| HDL-C < 40 mg/dL in men or HDL-C < 50 mg/dL in women | 3,421 (21.6) | 8,005 (26.8) | <.001 |
| SBP ≥ 130 mmHg or DBP ≥ 85 mmHg, | 5,834 (36.8) | 6,180 (20.7) | <.001 |
| WC ≥ 90 cm in men or WC ≥ 80 cm in women | 3,061 (19.3) | 8,468 (28.3) | <.001 |
| Lung function parameters |  |  |  |
| FEV1, mean (SD), L | 2.9 (0.8) | 2.0 (0.6) | <.001 |
| FVC, mean (SD), L | 3.6 (0.7) | 2.4 (0.5) | <.001 |
| FEV1, % predicted | 89.1 (22.0) | 83.8 (22.5) | <.001 |
| FVC, % predicted | 94.2 (15.2) | 91.0 (16.8) | <.001 |
| FEV1 /FVC ratio | 95.2 (20.8) | 92.5 (20.7) | <.001 |
| Lung function parameters, n (%) |  |  |  |
| FEV1 < 80% predicted | 4,084 (25.8) | 10,667 (35.6) | <.001 |
| FVC < 80% predicted | 2,348 (14.8) | 6,674 (22.3) | <.001 |
| Lung function diagnosis, n (%) |  |  |  |
| Normal ventilatory function | 11,855 (74.8) | 20,025 (66.9) | <.001 |
| Restrictive ventilatory defect | 2,058 (13.0) | 5,610 (18.7) |  |
| Obstructive ventilatory defect | 1,946 (12.3) | 4,294 (14.3) |  |

### 2. Effects of MetS on pulmonary function and ventilatory defect

The overall prevalence rate of MetS was estimated to be 15.9% in Taiwan, much higher in men than in women (18.6% versus 14.4%) (Table 1). Of these, 16.7% had RVDs and 13.6% had OVDs. Although higher proportion of male participants had MetS (18.6% vs 14.4%) and unhealthy lifestyle behaviors (current smoking, 22.0% vs 2.9%; current drinking, 13.1% vs 1.9%) than female participants in this study, rates of impaired pulmonary function were markedly higher among women for both types of ventilatory defects (RVD, 18.7% vs 13.0%; OVD, 14.3% vs 12.3%). In both sex strata, MetS increased risk of developing impaired pulmonary function FVC<80% (Odds Ratio [OR], 1.33-1.34), similar in magnitude to that of all participants (OR, 1.34), which suggested that the outcome was independent of sex. Similarly, the risks of MetS on developing impaired function FEV1<80%, predicted (OR, 1.19-1.23 vs 1.22) (Table 2) and RVD (OR, 1.31-1.36 vs 1.35) (Table 3) were both independent of sex. On the contrary, MetS did not increase the risk of developing OVD after covariate adjustment (Table 3). Table 4 shows subgroup analysis stratified by the MSS scores for the presence of metabolic abnormalities from none (MSS=0) to 5 (MSS=5). In general, both men and women were associated with increased risk of developing RVD from for MSS stratification of 2 to 5. Once again, the magnitude of risk for each MSS stratum for each sex was similar to that of all participant, which suggested that the risk of RVD was independent of sex. Table 5 compares the effects of individual metabolic abnormalities on the risk of RVD by age and sex categories. In general, both men and women with elevated BP (OR, 1.27-1.46 vs 1.33) and TGs (1.19 - 1.35 vs 1.22) abnormalities were more likely to develop RVD and the risks were independent of both sex and age strata. Conversely, sex-stratified subgroup analyses of the individual metabolic abnormalities indicated that only women with elevated FPG (OR, 1.14-1.38 vs 1.12) and men with abdominal obesity (OR, 1.24-1.30 vs 1.07) were more likely to develop RVD. Although the risk of RVD was associated with reduced HDL-C by sex (OR, 1.10-1.14) and for all participants (OR, 1.11), the risks of RVD were not statistically difference between age strata for men and only statically significance in women younger than 55 years (OR, 1.13).

**Table 2.**
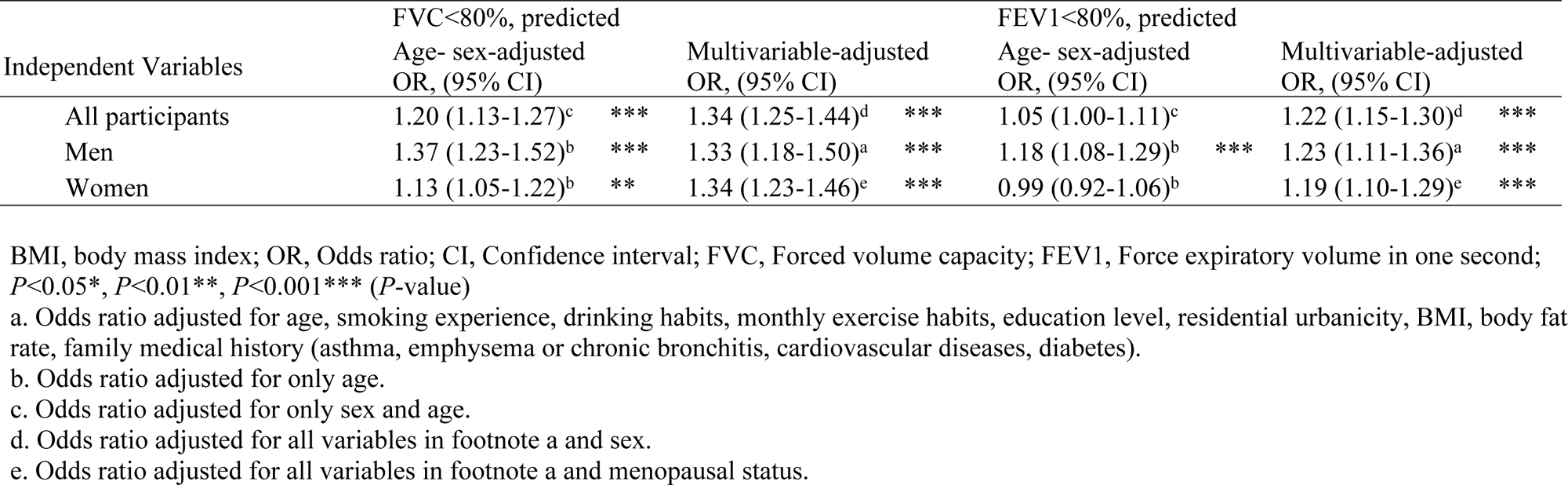
The effects of metabolic syndrome on the risk of impaired pulmonary function by sex.

**Table 3.** The effects of metabolic syndrome on the risk of ventilatory defect with sex.

| Independent Variables | Restrictive ventilatory defect |  |  |  | Obstructive ventilatory defect |  |  |
| --- | --- | --- | --- | --- | --- | --- | --- |
|  | Age- sex-adjusted<br>OR, (95% CI) |  | Multivariable-adjusted<br>OR, (95% CI) |  | Age- sex-adjusted<br>OR, (95% CI) |  | Multivariable-adjusted<br>OR, (95% CI) |
| All participants | 1.27 (1.19-1.35)<sup>c</sup> | *** | 1.35 (1.26-1.45)<sup>d</sup> | *** | 0.86 (0.79-0.93)<sup>c</sup> | *** | 0.97 (0.89-1.06)<sup>d</sup> |
| Men | 1.44 (1.29-1.61)<sup>b</sup> | *** | 1.31 (1.16-1.49)<sup>a</sup> | *** | 0.92 (0.81-1.04)<sup>b</sup> |  | 1.05 (0.91-1.21)<sup>a</sup> |
| Women | 1.20 (1.11-1.30)<sup>b</sup> | *** | 1.36 (1.24-1.48)<sup>e</sup> | *** | 0.82 (0.74-0.91)<sup>b</sup> | ** | 0.92 (0.82-1.03)<sup>e</sup> |

BMI, body mass index; OR, Odds ratio; CI, confidence interval; Restrictive ventilatory defect: FEV1/FVC ≥ 70% and FVC, predicted% < 80%; Obstructive ventilatory defect: FEV1/FVC < 70%; $P < 0.05^*$ , $P < 0.01^{**}$ , $P < 0.001^{***}$ ( $P$ -value)
a. Odds ratio adjusted for age, smoking experience, drinking habits, monthly exercise habits, education level, residential urbanicity, BMI, body fat rate, family medical history (asthma, emphysema or chronic bronchitis, cardiovascular diseases, diabetes).
b. Odds ratio adjusted for only age.
c. Odds ratio adjusted for only sex and age.
d. Odds ratio adjusted for all variables in footnote a and sex.
e. Odds ratio adjusted for all variables in footnote a and menopausal status.

**Table 4.** The effects of metabolic syndrome scores on the risk of restrictive ventilatory defect by sex.

| OR, (95% CI) | Non-metabolic syndrome |  |  | Metabolic syndrome |  |  |
| --- | --- | --- | --- | --- | --- | --- |
|  | MSS=0 | MSS=1 | MSS=2 | MSS=3 | MSS=4 | MSS=5 |
| Multivariable-adjusted<sup>b</sup> | 1 | 1.06 (0.99 -1.13) | 1.21 (1.11 -1.31) *** | 1.41 (1.28 -1.56) *** | 1.51 (1.33 -1.72) *** | 2.40 (1.94 -2.97) *** |
| Men<sup>a</sup> | 1 | 1.15 (1.00 -1.32) * | 1.34 (1.16 -1.56) *** | 1.52 (1.27 -1.81) *** | 1.58 (1.26 -1.99) *** | 2.75 (1.87 -4.04) *** |
| Women<sup>c</sup> | 1 | 1.04 (0.96 -1.13) | 1.16 (1.06 -1.28) ** | 1.38 (1.23 -1.55) *** | 1.49 (1.27 -1.74) *** | 2.26 (1.75 -2.91) *** |

BMI, body mass index; OR, Odds ratio; CI, confidence interval; MSS, metabolic syndrome score; $P<0.05^*$ , $P<0.01^{**}$ , $P<0.001^{***}$ ( $P$ -value)
a. Odds ratio adjusted for age, smoking experience, drinking habits, monthly exercise habits, education level, residential urbanicity, BMI, body fat rate, family medical history (asthma, emphysema or chronic bronchitis, cardiovascular diseases, diabetes).
b. Odds ratio adjusted for all variables in footnote a and sex.
c. Odds ratio adjusted for all variables in footnote a and menopausal status.

**Table 5.**
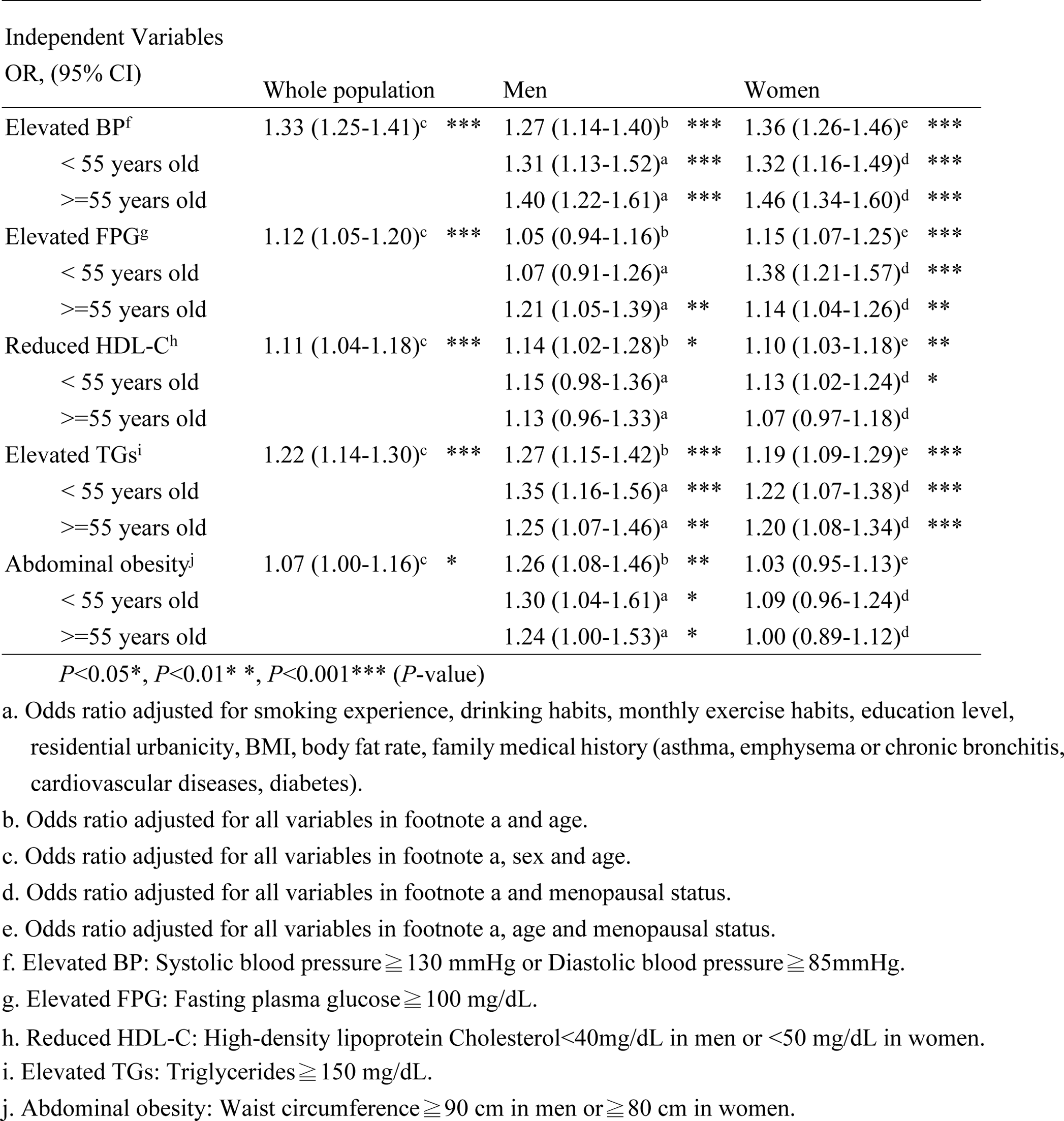
The effects of each metabolic component on the risk of restrictive ventilatory defect by sex and age.

| Independent Variables |  |  |  |  |  |  |
| --- | --- | --- | --- | --- | --- | --- |
| OR, (95% CI) | Whole population |  | Men |  | Women |  |
| Elevated BP<sup>f</sup> | 1.33 (1.25-1.41)<sup>c</sup> | *** | 1.27 (1.14-1.40)<sup>b</sup> | *** | 1.36 (1.26-1.46)<sup>e</sup> | *** |
| < 55 years old |  |  | 1.31 (1.13-1.52)<sup>a</sup> | *** | 1.32 (1.16-1.49)<sup>d</sup> | *** |
| ≥55 years old |  |  | 1.40 (1.22-1.61)<sup>a</sup> | *** | 1.46 (1.34-1.60)<sup>d</sup> | *** |
| Elevated FPG<sup>g</sup> | 1.12 (1.05-1.20)<sup>c</sup> | *** | 1.05 (0.94-1.16)<sup>b</sup> |  | 1.15 (1.07-1.25)<sup>e</sup> | *** |
| < 55 years old |  |  | 1.07 (0.91-1.26)<sup>a</sup> |  | 1.38 (1.21-1.57)<sup>d</sup> | *** |
| ≥55 years old |  |  | 1.21 (1.05-1.39)<sup>a</sup> | ** | 1.14 (1.04-1.26)<sup>d</sup> | ** |
| Reduced HDL-C<sup>h</sup> | 1.11 (1.04-1.18)<sup>c</sup> | *** | 1.14 (1.02-1.28)<sup>b</sup> | * | 1.10 (1.03-1.18)<sup>e</sup> | ** |
| < 55 years old |  |  | 1.15 (0.98-1.36)<sup>a</sup> |  | 1.13 (1.02-1.24)<sup>d</sup> | * |
| ≥55 years old |  |  | 1.13 (0.96-1.33)<sup>a</sup> |  | 1.07 (0.97-1.18)<sup>d</sup> |  |
| Elevated TG<sup>i</sup> | 1.22 (1.14-1.30)<sup>c</sup> | *** | 1.27 (1.15-1.42)<sup>b</sup> | *** | 1.19 (1.09-1.29)<sup>e</sup> | *** |
| < 55 years old |  |  | 1.35 (1.16-1.56)<sup>a</sup> | *** | 1.22 (1.07-1.38)<sup>d</sup> | *** |
| ≥55 years old |  |  | 1.25 (1.07-1.46)<sup>a</sup> | ** | 1.20 (1.08-1.34)<sup>d</sup> | *** |
| Abdominal obesity<sup>j</sup> | 1.07 (1.00-1.16)<sup>c</sup> | * | 1.26 (1.08-1.46)<sup>b</sup> | ** | 1.03 (0.95-1.13)<sup>e</sup> |  |
| < 55 years old |  |  | 1.30 (1.04-1.61)<sup>a</sup> | * | 1.09 (0.96-1.24)<sup>d</sup> |  |
| ≥55 years old |  |  | 1.24 (1.00-1.53)<sup>a</sup> | * | 1.00 (0.89-1.12)<sup>d</sup> |  |

$P < 0.05^*$ , $P < 0.01^{**}$ , $P < 0.001^{***}$ ( $P$ -value)
- Odds ratio adjusted for smoking experience, drinking habits, monthly exercise habits, education level, residential urbanicity, BMI, body fat rate, family medical history (asthma, emphysema or chronic bronchitis, cardiovascular diseases, diabetes). - Odds ratio adjusted for all variables in footnote a and age. - Odds ratio adjusted for all variables in footnote a, sex and age. - Odds ratio adjusted for all variables in footnote a and menopausal status. - Odds ratio adjusted for all variables in footnote a, age and menopausal status. - Elevated BP: Systolic blood pressure $\geq 130$ mmHg or Diastolic blood pressure $\geq 85$ mmHg. - Elevated FPG: Fasting plasma glucose $\geq 100$ mg/dL. - Reduced HDL-C: High-density lipoprotein Cholesterol $< 40$ mg/dL in men or $< 50$ mg/dL in women. - Elevated TGs: Triglycerides $\geq 150$ mg/dL. - Abdominal obesity: Waist circumference $\geq 90$ cm in men or $\geq 80$ cm in women.

## Discussion

Our study evidence suggested that men and women aged 30 years or older with MetS were more likely to suffer from impaired pulmonary function and increased risk of developing RVD, and its risk increased with increasing numbers of metabolic abnormalities. Despite much higher prevalence of MetS and unhealthy lifestyle (smoking and drinking) behaviors in men than women, similar rates of increased risk of developing RVD (31%-36%) were found in both sexes, whereas the difference in the risk of OVD was not significant after adjusting for baseline demographic and lifestyle behaviors. Additionally, elevated BPs and TGs were both important predictors of RVD. In contrast, only men with abdominal obesity (or increasing WC) and women with dysglycemia (elevating fasting blood glucose) were more likely to develop RVD. Many studies demonstrated that the risk of developing RVD was associated with MetS [7, 19–24]. We observed an increased risk of RVD with increasing numbers of metabolic abnormalities, which was consistent with those reported by other researchers [7], [19, 25]. However, the results are not consistent in terms of the correlation between MetS and OVD [26]. Lam et al. classified patients of COPD from I to IV according to GOLD criteria, and found only a significant correlation between MetS and the worst COPD groups [27], which came up with similar results in a study on Japanese men [28]. In another study, Lee et al. study observed that diabetes is positively associated with RVD (OR:2.025, 95% CI:1.264-3.244), but no correlative with OVD (OR:0.982, 95%CI:0.634-1.519). They also found that subjects with RVD had the highest hs-CRP than other respiratory dysfunction [29].

Abdominal obesity is recognized as a major predicting factor for impaired respiratory function, which has been confirmed in many studies [25, 30]. Wang et al. found that abdominal obesity lowers FVC and FEV1 values but it did not affect the FEV1/FVC value [25]. Our study shows that men’s WC over 90 cm would decrease ventilatory function, such as FVC<80% predicted and FEV1<80% predicted. In contrast, in a study of 6,945 Taiwanese participants, a negative correlation was reported between waist circumference and FVC % predicted in men younger than 45 years while no correlation between metabolic abnormality and FVC % predicted was observed in women [7]. On the other hand, the Korean National Health and Nutrition Examination Survey in 2007 observed that WC, BP, FPG, and HDL-C independently related to FVC % of predicted in men while WC significantly correlated with FVC predicted (%) in women [6]. Obese men present as the deposit of fat in the chest, abdomen, and visceral organs, known as central obesity. In women, fat is usually stored in the buttocks, thighs, hips, and subcutaneous tissue, known as peripheral obesity [31]. Accumulated fat in the mediastinum and abdomen decreases the compliance of the chest wall, causing breathing pattern changes. Intra-abdominal pressure and intrapleural pressure would slightly increase in the obese which decreases differential pressure from the atmosphere to the chest leading to a ventilatory flow limit. In addition, the downward movement of the diaphragm and expansion of the chest would be restricted by adipose tissue in the abdomen, decreasing expiratory reserved volume and functional residual capacity making work of breath increasing and ventilatory efficiency lowering [15, 31]. However, it is still unclear whether obesity lower the compliance of lung, chest, or both [32]. Further studies are needed to confirm the association.

MetS is a chronic inflammatory state that would trigger immune response and proliferate inflammatory substances to reduce the pulmonary function. In our study, we found that men with abdominal obesity (or increasing WC) and women having dysglycemia (elevating FPG) had a higher risk of respiratory function (RVD) than those without these metabolic abnormalities. This indicated a sex-specific association between abnormal fasting blood glucose and the secretion of inflammatory substances. Kawamoto et al. reported a significant sex-specific correlation between fasting blood glucose and hsCRP, and women had a large increasing amplitude than men [33]. Elevating hs-CRP amplitude different explained that women with high blood glucose will have a higher risk of cardiovascular disease than men in the future [34]. Likewise, Adipocyte accumulation due to obesity will secrete adipocytokines, such as Interleukin-6, TNF-α, interferon-γ, and leptin, and stimulate the bronchial smooth muscle contraction and induce macrophages to move into the alveoli provoking chronic inflammation damage alveoli and airway, and increase respiratory symptoms [35].

There are some limitations to be considered while interpreting our study results. First, the measurement of the respiratory function with spirometry, rather than using the more bulky and expensive equipment such as body plethysmography, did not provide data on total lung capacity and residual volume, which could have resulted in residual confounding. Nevertheless, the use of spirometry with properly trained technicians is sufficient for early screening of respiratory function in the community. Second, respiratory symptoms that may be associated with ventilatory defect, such as cough and dyspnea, were not available in the Taiwan Biobank. We were therefore unable to assess the relationship between MetS and respiratory symptoms. Finally, the lack of data on inflammatory substances such as hs-CPR prevent further investigation on the association between inflammatory substances and impaired respiratory function.

## Conclusion

Despite limitation, our study evidences suggested that metabolic syndromes were important predictors of impaired pulmonary function and increased risk of developing restrictive ventilatory defects, and its risk increased with increasing numbers of metabolic abnormalities. If gender-specific prevention strategies can be tailored for individuals at increased risk of impairment pulmonary function, further studies are required to determine the metabolic effects of medications as well as modifiable lifestyle behaviors on metabolic abnormalities

## Author Contributions

**Conceptualization:** Ya-Chun Chu; Chi-Chiang Yang; Hsin-Hung Wu; Jerry Cheng-Yen Lai.

**Data curation:** Ya-Chun Chu; Shaw-Ji Chen; Jerry Cheng-Yen Lai.

**Formal analysis:** Ya-Chun Chu; Chi-Chiang Yang; Shaw-Ji Chen; Jerry Cheng-Yen Lai.

**Funding acquisition:** Jerry Cheng-Yen Lai; Pei-Ling Cheng; Mei-Chuan Wu.

**Investigation:** Ya-Chun Chu; Pei-Ling Cheng.

**Methodology:** Ya-Chun Chu; Shaw-Ji Chen; Hsin-Hung Wu; Jerry Cheng-Yen Lai.

**Project administration:** Chi-Chiang Yang; Pei-Ling Cheng; Hsin-Hung Wu.

**Resources:** Chi-Chiang Yang; Pei-Ling Cheng; Mei-Chuan Wu.

**Software:** Pei-Ling Cheng; Jerry Cheng-Yen Lai.

**Supervision:** Shaw-Ji Chen; Mei-Chuan Wu; Hsin-Hung Wu; Jerry Cheng-Yen Lai.

**Validation:** Ya-Chun Chu; Hsin-Hung Wu; Jerry Cheng-Yen Lai.

**Visualization:** Ya-Chun Chu; Chi-Chiang Yang; Hsin-Hung Wu.

**Writing - orginal draft:** Ya-Chun Chu; Chi-Chiang Yang; Shaw-Ji Chen; Jerry Cheng-Yen Lai.

**Writing - review&editing:** Ya-Chun Chu; Hsin-Hung Wu; Jerry Cheng-Yen Lai.

## Data availability statement

The data that support the findings of this study are available from the Taiwan Biobank but restrictions apply to the availability of these data, which were used under license for this study. Data are however available from the authors upon reasonable request with the permission of Taiwan Biobank.

